# Evaluation of peripheral enhancement on contrast-enhanced CT and corresponding pathological findings in colorectal liver metastases after preoperative chemotherapy

**DOI:** 10.1101/2021.10.27.21265582

**Authors:** Akio Tamura, Kazuyuki Ishida, Misato Sone, Kunihiro Yoshioka

**Affiliations:** Department of Radiology, Iwate Medical University School of Medicine, 2-1-1 Idaidori, Yahaba-cho, Shiwa-gun, Iwate Prefecture 028-3695, Japan; Department of Diagnostic Pathology, Dokkyo Medical University, 880 Kitakobayashi, Mibu, Shimotsuga, Tochigi, Japan

## Abstract

**Objective:** To correlate peripheral enhancement on contrast-enhanced computed tomography (CE-CT) of post-chemotherapy colorectal liver metastases (CRLM) patients with the pathological findings.

**Methods:** Forty-four patients with CRLM who underwent hepatic resection after preoperative chemotherapy between 2008 and 2013 were included. Two radiologists blinded to the histopathology findings performed a consensus categorization of the marginal contrast effects of CRLM on CE-CT as follows: Group 1, smooth margin without enhancement; Group 2, smooth margin with an enhanced rim; and Group 3, fuzzy margin with/without an enhanced rim. The Kruskal-Wallis test was used to compare the imaging findings with the histological findings.

**Results:** The percentage of infarct-like necrosis was significantly higher in CRLM with smooth margins than in those with fuzzy margins (p<0.001, r=0.62). The percentage of viable cells was lowest in CRLM with smooth margins without enhancement (p<0.001, r=0.60).

**Conclusions:** Our findings suggest that the type of necrosis is related to the nature of the margins, and the presence of residual cells are related to peripheral enhancement.

**Advances in knowledge:** In CRLM, following chemotherapy, the presence of residual cells and dangerous haloes is related to the contrast effect of the tumor margins, suggesting that tumor angiogenesis affects the contrast effect.

## Introduction

The progress in chemotherapy for advanced colorectal cancer has been remarkable, from the era of 5-FU alone to the present, in which FOLFIRI and FOLFOX in combination with irinotecan and oxaliplatin have been established as primary and secondary therapies with the introduction of molecular targeted agents.^1–3^ With the widespread use of highly effective drugs, including molecularly targeted agents, there has been an increase in the number of conversions of unresectable cases of colorectal liver metastases (CRLM) to resectable ones, and there have been more opportunities to compare the imaging and histological findings of CRLM after chemotherapy. In daily clinical practice, response to chemotherapy is determined by diagnostic imaging, especially contrast-enhanced computed tomography (CE-CT). Chun et al. proposed the CT morphologic criterion, noting that CRLM after chemotherapy show changes in the homogeneity of intratumor concentration, marginal characteristics, and contrast effect of the margins, even if the tumor size is unchanged.^4^ The CT morphologic criteria have been reported to correlate with pathologic response rates and patient survival rates, but few studies have specified the corresponding histological findings that reflect these changes.^4–6^ Peripheral enhancement is a characteristic finding of CRLM, and in recent years, peripheral enhancement has been considered an important indicator in determining the response to treatment of CRLM after chemotherapy.^4,7–10^ The prevalence of peripheral enhancement, however, varies widely among studies, and what peripheral enhancement appears as histologically has not been determined. This difference in perception may lead to different outcomes in determining the treatment efficacy. In this study, we attempted to directly correlate the findings of peripheral enhancement on CE-CT of post-chemotherapy CRLM patients with their pathological findings to better understand peripheral enhancement.

## Methods and materials

This retrospective study was approved by the institutional review board, and informed consent waiver was obtained. In this retrospective study, patients with CRLM who underwent hepatic resection for CRLM after preoperative chemotherapy between 2008 and 2013 were included. The inclusion criteria were the performance of preoperative dynamic-enhanced CT within 2 months before surgical resection, detectable CRLM on preoperative CT, and a confirmed diagnosis of CRLM based on histopathologic examination.

All examinations were performed with similar imaging parameters, except for the CT system (Light Speed VCT 64 Slice CT, GE Healthcare, Wauwatosa, WI, USA; Toshiba Aquilion 64, Toshiba Medical Systems, Otawara, Japan). Iohexol (Omnipaque 300; Daiichi-Sankyo, Tokyo, Japan), iopamidol (Iopamiron 300; Nihon Schering, Osaka, Japan), or iomeprol (Iomeron 350; Eisai, Tokyo, Japan), was administered into the antecubital vein at a dose of 600 mgI/kg). The scan delays for arterial and portal venous phase imaging were determined using an automatic bolus-tracking program (Smart Prep software: GE Healthcare, Wauwatosa, WI, USA; Real Prep: Toshiba Medical Systems, Otawara, Japan). The region of interest (ROI) cursor was placed in the aorta at the level of the diaphragmatic dome; scanning for the arterial and portal venous phases began automatically at 20 s and 60 s, respectively, after the trigger threshold of 100 Hounsfield Units was reached. However, the arterial phase images were not evaluated in this study. Images were reconstructed using soft tissue window settings, with a 3 mm slice thickness and interval.

Two radiologists (M.S. and A.T. with 4 and 13 years’ experience in abdominal CT, respectively) who were blinded to the histopathology findings performed a consensus categorization of the marginal contrast effects of CRLM on CE-CT (portal venous phase), as follows: Group 1, smooth margin without enhancement; Group 2, smooth margin with an enhanced rim; and Group 3, fuzzy margin with/without an enhanced rim.

Liver resection specimens of CRLM after chemotherapy were stained with hematoxylin and eosin, and the following findings were evaluated: the percentage of dangerous halos in the tumor circumference, percentage of fibrosis in the tumor, percentage of infarct-like necrosis (ILN), and percentage of residual tumor cells. In patients with multiple metastases, the lesion with the greatest diameter, which fit a radiological target lesion, was examined. Dangerous halos indicate that the surviving tumor cells have infiltrated the surrounding liver parenchyma, and there is no fibroinflammatory response.^11^ ILN was defined as a large confluent area of eosinophilic cytoplasmic debris located in the center of the lesion, with no or minimal intermingled nuclear debris.^12^ Usual necrosis, defined as patchy distribution of nuclear debris, confluent necrosis, and borderline necrosis, is the opposite of ILN, but was not directly assessed in this study.^12^ Slides were evaluated by two pathologists (with 14 and 20 years of experience in pathology, respectively) blinded to the subjects’ clinical data.

The Kruskal-Wallis test was used to compare the three groups. If there was a significant difference (p<0.05) among the three groups, post-hoc analysis by the Bonferroni test was performed. In addition, the Mann–Whitney U test was used to investigate the effect size: r > 0.1, small effect; > 0.3, medium effect; and > 0.5, large effect. Statistical analysis was performed using the SPSS software (version 24; IBM Corp., Armonk, NY).

## Results

We examined 44 patients (30 males, age range, 41-79 years; mean age, 64.4 years) who had undergone the first hepatic resection for CRLM following chemotherapy (23 patients received FOLFOX or FOLFIRI with bevacizumab, 9 patients received FOLFOX or FOLFIRI, and 11 patients received 5-FU only). Correlations between peripheral enhancement on contrast-enhanced CT and histopathological findings are shown in Table 1. The percentage of dangerous haloes was not significantly different among the three groups (p=0.169), but that in Group 1 tended to be lower than that in the other groups. The percentage of ILN was significantly higher in Groups 1 and 2, which had smooth margins, than in Group 3 (p<0.001, r=0.62). The percentage of viable cells was the lowest in tumors with smooth margins without enhancement, followed by Groups 2 and 3, with a significant difference between the groups (p<0.001, r=0.60). A typical case is shown in Figure 1.

**Table 1.**
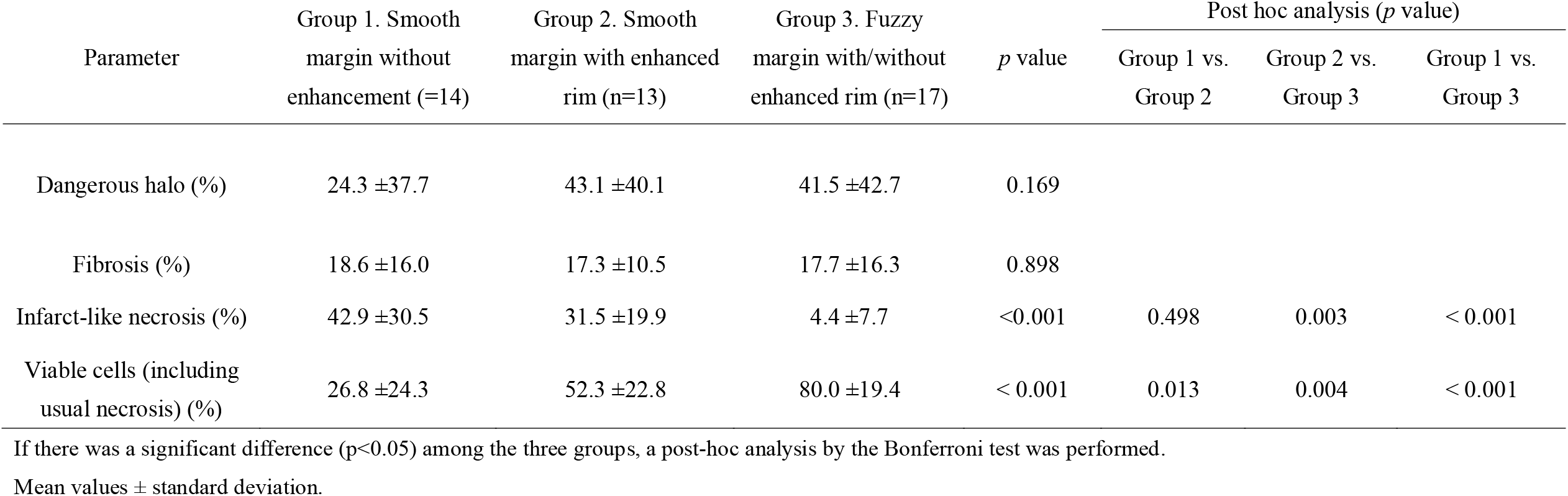
Correlations between peripheral enhancement on contrast-enhanced CT and histopathological findings

| Parameter | Group 1. Smooth<br>margin without<br>enhancement (=14) | Group 2. Smooth<br>margin with enhanced<br>rim (n=13) | Group 3. Fuzzy<br>margin with/without<br>enhanced rim (n=17) | *p* value | Post hoc analysis (*p* value) |  |  |
| --- | --- | --- | --- | --- | --- | --- | --- |
|  |  |  |  |  | Group 1 vs.<br>Group 2 | Group 2 vs.<br>Group 3 | Group 1 vs.<br>Group 3 |
| Dangerous halo (%) | 24.3 ±37.7 | 43.1 ±40.1 | 41.5 ±42.7 | 0.169 |  |  |  |
| Fibrosis (%) | 18.6 ±16.0 | 17.3 ±10.5 | 17.7 ±16.3 | 0.898 |  |  |  |
| Infarct-like necrosis (%) | 42.9 ±30.5 | 31.5 ±19.9 | 4.4 ±7.7 | <0.001 | 0.498 | 0.003 | < 0.001 |
| Viable cells (including<br>usual necrosis) (%) | 26.8 ±24.3 | 52.3 ±22.8 | 80.0 ±19.4 | < 0.001 | 0.013 | 0.004 | < 0.001 |

If there was a significant difference ( $p<0.05$ ) among the three groups, a post-hoc analysis by the Bonferroni test was performed.
Mean values ± standard deviation.

**Fig 1.**
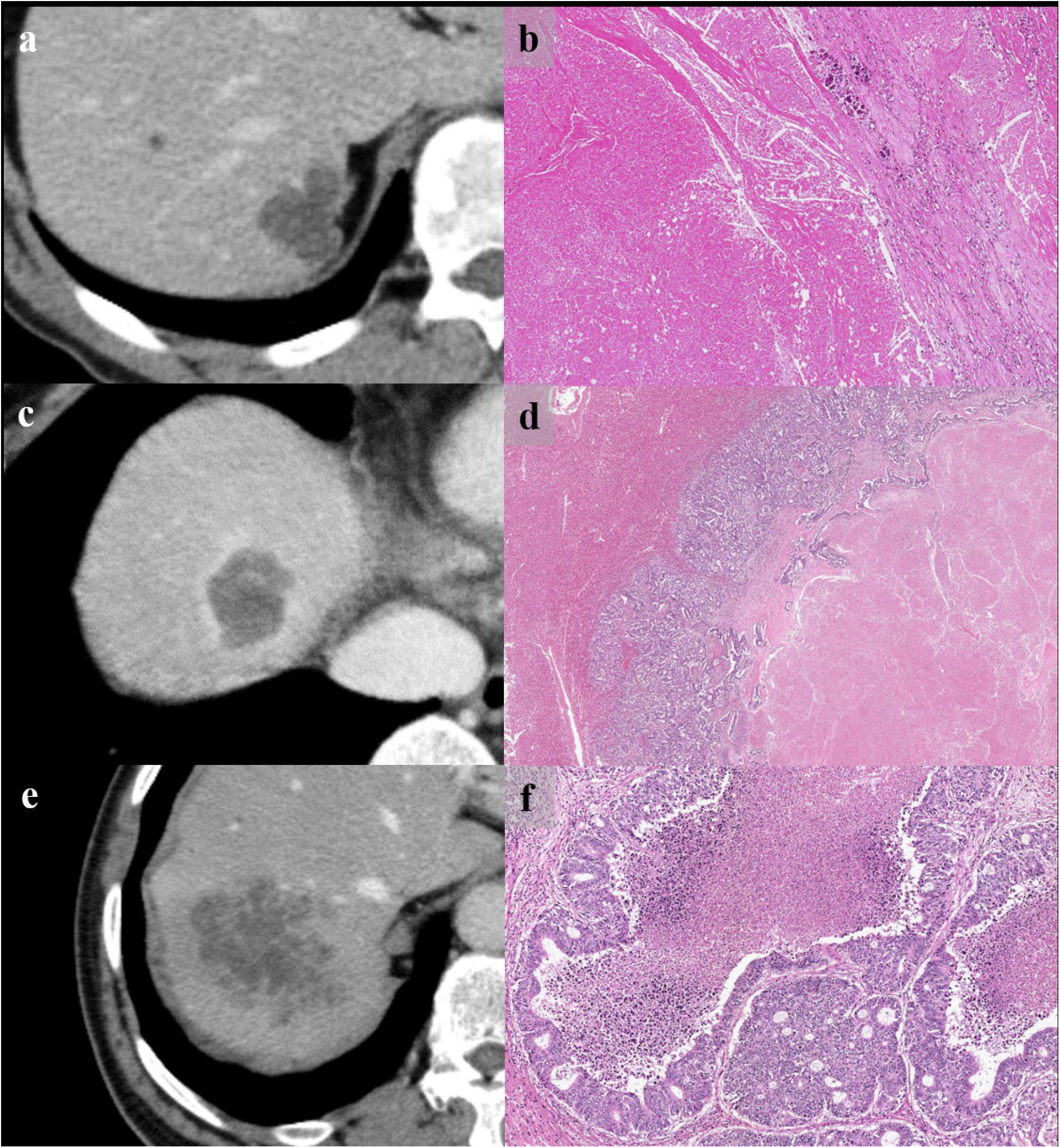
CE-CT images and H and E-stained sections showing different histologic patterns. a Group 1: CE-CT showing tumors with smooth margins and resolved contrast effects in the portal venous phase. b A high magnification view of the HE-stained section shows infarct-like necrosis with large confluent areas that lack nuclear debris. A rim of fibrosis surrounds the necrosis. No residual tumor is seen, and it a pathological complete response is observed. c Group 2: CE-CT shows colorectal liver metastasis with enhanced rims and well-defined margins at the portal venous phase. d Microscopically, the center of the tumor shows infarct-like necrosis. There is residual tumor at the margin of the tumor, and the overall morphology of the tumor presents as a dangerous halo. e Group 3: Colorectal liver metastasis shows a fuzzy margin without an enhanced rim. f Microscopically, the center of the tumor shows usual necrosis containing nuclear debris and is surrounded by viable tumor cells. CE-CT, contrast-enhanced computed tomography; HE, hematoxylin and eosin

## Discussion

Our results showed that when ILN was dominant and few remaining cells were present, the margins were clear and there was no contrast effect, and when ILN was dominant and abundant remaining cells were present, the margins were clear and there was a contrast effect at the tumor margins. The type of necrosis is related to the nature of the margins, and the presence of residual cells and tumor angiogenesis are related to the contrast effect of the tumor margins. CRLM often present with a heterogeneous appearance, reflecting the presence of fibrosing necrosis spreading from the center of the tumor and tumor cells spreading around it. The tumor content tends to be homogenous hypoattenuation after chemotherapy, especially after chemotherapy with bevacizumab. This finding is histologically reflective of ILN, and uniformly distributed ILN may be reflected by uniform hypoattenuation within the tumor on CT images.^13^ On the other hand, the heterogeneous appearance within the tumor is thought to reflect the predominance of UN. ILN has been reported to be observed in lesions that responded well to chemotherapy, especially more frequently in cases treated with bevacizumab.^13,14^ Chang et al. reported that patients with ILN had significantly longer disease-free survival than those with UN.^12^ CRLM are often visualized as lesions with fuzzy tumor margins on CT. However, as the necrosis inside the tumor progresses after chemotherapy, the contrast with the normal liver parenchyma becomes relatively clear, and the margins of the tumor become easier to recognize. Various interpretations of the imaging findings corresponding to the contrast effect at the tumor margins have been reported, including desmoplastic reaction, inflammatory cell infiltration, and proliferation of residual tumor cells.^7–10^ The incidence of these findings varies greatly among studies. Inaba et al. compared the contrast patterns of liver metastases from CRLM in CT angiography with postoperative pathological specimens and reported that lesions with dense tumor margins showed extensive necrosis or fibrosis in the center of the tumor surrounded by tumor cells arranged in a ring.^10^ They also reported that lesions with heterogeneously contrasted tumors had dense tumor cells at the margins but also showed mixed fibrosis and necrosis in the center. This report was probably based on a specimen resected without preoperative chemotherapy. In this study, we investigated the relationship between the contrast effect on the margins, tumor cells, and type of necrosis in CRLM after preoperative chemotherapy. As mentioned above, when ILN is dominant and there are few residual cells after preoperative chemotherapy, the border of the tumor edge is clear and there is no contrast effect, while when ILN is dominant and there are many residual cells, there is a contrast effect on the tumor edge. The type of necrosis is related to the morphology of the tumor margins, and the presence or absence of residual cells and dangerous haloes is related to the contrast effect of the tumor margins, suggesting that tumor angiogenesis affects the contrast effect. Based on Inaba et al. and our study, it should be noted that evaluation of the contrast effect of the tumor margins in CT is focused on their contrast with the surrounding tissue and is not an absolute evaluation, especially considering that it involves visual inspection. Therefore, it may be greatly affected by the status of tumor necrosis.

The limitation of this study was the small number of cases, which prevented us from further subdividing the groups for more detailed studies. The percentage of dangerous haloes was not significantly different among the three groups. A greater sample size could help obtain significant results, and therefore, further studies are recommended.

## Conclusion

The findings of this study suggest that in CRLM, contrast enhancement of the tumor margins reflects the presence of residual cells, but the contrast may be affected by the nature of the internal necrosis.

## Data Availability

All data produced in the present work are contained in the manuscript

## Acknowledgments

The authors would like to thank Dr. Noriyuki Uesugi from Department of Molecular Diagnostic Pathology, Iwate Medical University.

